# ULCERATIVE COLITIS-DERIVED COLONOID CULTURE: A Multi-Mineral-Approach to Improve Barrier Protein Expression

**DOI:** 10.1101/2019.12.12.19014662

**Authors:** Muhammad N Aslam, Shannon D McClintock, Durga Attili, Shailja Pandya, Humza Rehman, Daniyal M Nadeem, Mohamed Ali H Jawad-Makki, Areeba H Rizvi, Maliha M Berner, Michael K Dame, Danielle Kim Turgeon, James Varani

## Abstract

**Background:** Recent studies demonstrated that Aquamin^®^, a calcium-, magnesium-, and multiple trace element-rich natural product, improves barrier structure and function in colonoids obtained from tissue of healthy subjects. The goal of the present study was to determine if the colonic barrier could be improved in tissue from subjects with ulcerative colitis (UC).

**Methods:** Colonoid cultures were established with tissue from 9 individuals with UC. The colonoids were then incubated for a 2-week period under control conditions (i.e., in culture medium with a final calcium concentration of 0.25 mM) or in the same medium supplemented with Aquamin^®^ to provide 1.5 – 4.5 mM calcium. Effects on differentiation and barrier protein expression were determined using several approaches: phase-contrast & scanning electron microscopy, quantitative histology & immunohistology, mass spectrometry-based proteome assessment and transmission electron microscopy.

**Results:** Aquamin^®^-treated colonoids demonstrated a modest up-regulation of tight junctional proteins but stronger induction of adherens junction proteins and desmosomal proteins. Increased desmosomes were seen at the ultrastructural level. Proteomic analysis also demonstrated increased expression of basement membrane proteins and hemidesmosomal components. Proteins expressed at the apical surface (mucins and trefoils) were also increased as were several additional proteins with anti-microbial activity or that modulate inflammation.

**Conclusion:** A majority of individuals including patients with UC do not reach the recommended daily intake for calcium and other minerals. The findings presented here suggest that adequate mineral intake might improve the colonic barrier.

## INTRODUCTION

An intact colonic barrier is necessary for gastrointestinal health (1). Colonic barrier dysfunction is a consistent feature of inflammatory bowel disease, seen in both Crohn’s disease and ulcerative colitis (UC) (2-5). Barrier dysfunction is also seen in irritable bowel syndrome (6,7) and celiac disease (7), as well as in conjunction with acute bacterial infection of the gastrointestinal tract (8). While barrier dysfunction can result from toxic insults or from inflammatory attack on the epithelial cells lining the colon, it is now becoming recognized that pre-existing weaknesses in the gastrointestinal barrier may predispose the tissue to inflammation and injury.

Environmental factors, especially diet, have a significant impact on colonic barrier health. A high-fat diet has been shown to increase intestinal permeability (8) as has hyperglycemia (9). Most studies have focused on tight junctional complexes as the primary regulator of permeability, and high-fat or high-sugar intake has been shown to affect these structures (8,9). Our own work has demonstrated the importance of calcium as a regulator of the colonic barrier. In a recent study (10) we showed that increasing the extracellular calcium concentration from a basal level of 0.25 mM to as high as 3.0 mM in human colon organoid (colonoid) culture had a modest effect on expression of tight junction proteins, but dramatically up-regulated desmosomal proteins (desmoglein-2, desmocollin-2, and desmoplakin). This was accompanied by an actual increase in desmosomes seen at the ultrastructural level. A substantial increase in multiple basement membrane components including laminin α, β and γ chains as well as nidogen and perlecan (basement membrane heparin sulfate proteoglycan) was also observed in calcium-treated colonoids.

As part of our investigation, we carried out assays to assess the effects of calcium-supplementation on trans-epithelial electrical resistance (TEER) and colonoid cohesion (11). Consistent with tight junctional protein expression under control conditions, TEER values were high to begin with. In spite of the high background, electrical resistance still increased in response to calcium-supplementation. Colonoid cohesion, consistent with the dramatic increase in desmosomes and basement membrane proteins in response to intervention, was low in basal medium but increased substantially in treated colonoids. Of interest, while calcium provided as a single agent was effective, a calcium- and magnesium-rich multi-mineral natural product (Aquamin^®^) was more effective than calcium alone.

These findings indicate that colonoid culture technology can provide a useful tool for elucidating factors that affect barrier structure and function in the colon. The present study extends this work. Here we describe results with colonoid cultures established with tissue from UC patients. The UC-derived colonoids were maintained under control conditions (0.25 mM calcium) or treated with Aquamin^®^ - the same multi-mineral natural used in our previous studies - at amounts bringing the final calcium level to 1.5 - 4.5 mM. Effects on components of the barrier were determined using several approaches.

## MATERIALS AND METHODS

Aquamin^®^. Aquamin^®^ is a multi-mineral natural product obtained from the mineralized remains of red marine algae of the *Lithothamnion* family. Aquamin^®^-Soluble, the form used here, has been used in our previous colonoid studies (10-12). It consists of calcium (approximately 12% by weight), a calcium to magnesium ratio of 12:1 and detectable levels of 72 other trace minerals – essentially all of the minerals accumulated in the algae fronds from seawater. During growth, the mineralized fronds break off from the viable algae and fall to the ocean floor from where they are harvested. After harvest, the fronds are processed to remove organic material, leaving only the mineral remains. Aquamin^®^ is sold as a food supplement (GRAS 00028) and is used in various products for human consumption in Europe, Asia, Australia, and North America [Marigot Ltd, Cork, IR]. A single batch of Aquamin^®^ was used in the present study. Mineral analysis of this batch was provided by an independent laboratory (Advanced Laboratories; Salt Lake City, Utah) using Inductively Coupled Plasma Optical Emission Spectrometry. Mineral composition is provided in Supplement Table 1.

### Establishment of colonoid cultures from UC biopsies

De-identified colonic tissue (2.5-mm biopsies) was obtained from the sigmoid colon of eleven subjects during colonoscopy. The study was approved by the Institutional Review board (IRBMED) at the University of Michigan and all subjects provided written informed consent prior to endoscopy.

Colonoid cultures were established as described in our recent reports with tissue from healthy subjects (10-13). Briefly, biopsies were finely minced on ice using a #21 scalpel and seeded into matrigel (Corning), prepared at 8 mg/mL. A size of 40 μm (diameter) was considered ideal for tissue fragment size. During the expansion phase (between two and five weeks), colonoids were incubated in L-WRN medium containing 10% fetal bovine serum, and providing a source of Wnt3a, R-spondin-3 and Noggin. The medium was supplemented with small molecule inhibitors: 500 nM A 83-01 (Tocris), a TGF-β inhibitor, 10 µM SB 202190 (Sigma), a p38 inhibitor and 10 µM Y27632 (Tocris) as a ROCK inhibitor. For the first 10 days of culture, the medium was also supplemented with 2.5 µM CHIR9902 (Tocris) to enhance Wnt signaling.

After establishment, experiments were carried out in a mixture of L-WRN culture medium diluted 1:4 with KGM Gold, a serum-free, calcium-free culture medium (Lonza) designed for epithelial cell proliferation. The final serum concentration in the media was 2.5% and final calcium concentration was 0.25 mM. This “control treatment medium” was compared to the same medium supplemented with Aquamin^®^ to provide a final calcium concentration of 1.5, 2.1, 3.0 or 4.5 mM. Fresh medium and treatments were provided at 2-day intervals. The last 2-day medium collected before harvest at day-14 were saved for cytokine assessment. At the end of the in-life phase, colonoid samples were evaluated for gross appearance using both phase-contrast microscopy and scanning electron microscopy (SEM). Other samples were prepared for histology and examined at the light-microscopic level after staining with hematoxylin and eosin. The same preparation was used for immunohistology. Additional samples were fixed in glutaraldehyde and used for transmission electron microscopy (TEM). Finally, samples were frozen in liquid nitrogen and allocated to proteomic analysis.

### Phase-contrast microscopy

Phase-contrast microscopy (Hoffman Modulation Contrast – Olympus IX70 with a DP71 digital camera) was used to assess colonoids in whole-mount for change in size and shape.

### Histology, immunohistology and Morphometric analysis

At the end of the in-culture phase, colonoids were isolated from Matrigel using 2mM EDTA and fixed in 10% formalin for 1 hour. Fixed colonoids were suspended in HistoGel (Thermo Scientific) and then processed for histology (i.e., hematoxylin and eosin staining) or for immunohistology. For immunostaining, freshly-cut sections (5-6 microns) were rehydrated and subjected to heat-induced epitope retrieval with high pH or low pH FLEX TRS Retrieval buffer (Agilent Technologies, 154 #K8004; Santa Clara, CA) for 20 minutes. After peroxidase blocking, antibodies were applied at appropriate dilutions at room temperature for 30 or 60 minutes depending on the manufacturer’s recommendation. The FLEX HRP EnVision System (Agilent Technologies) was used for detection with a 10-minute DAB chromagen application. Supplement Table 2 provides a list of antibodies used, their source and additional experimental details.

The sections of immunostained colonoid tissue on glass slides were digitized using the Aperio AT2 whole slide scanner (Leica Biosystems) at a resolution of 0.5µm per pixel with 20X objective. These scanned images were housed on a server and accessed using Leica Aperio eSlide Manager (Version 12.3.2.5030), a digital pathology management software. The digitized histological sections were viewed and analyzed using Aperio ImageScope (Version 12.4.0.5043 by Leica Biosystems), a slide viewing software. Brightfield Immunohistochemistry Image Analysis tools (Leica) were used to quantify immunostains used in this study. Aperio Nuclear Algorithm (v9) was used for proliferation marker (Ki67) quantification. This algorithm measures intensity of nuclear staining and separates those into very intense to no nuclear staining (3+, 2+, 1+ and 0 respectively). Nuclei with higher intensity (3+ and 2+) were used here for comparison. The Aperio Positive Pixel Count Algorithm (v9) was used to quantify expression of the other markers. It quantifies number and the intensity of pixels of a specific color in a digitized image. Positivity was calculated with respective numbers of strong positive and positive pixels against total pixels.

### SEM and TEM

Colonoid specimens were fixed *in situ* in 2.5 percent glutaraldehyde in 0.1 M Sorensen’s buffer, pH 7.4, overnight at 4°C. After subsequent processing for SEM or TEM as described previously (14), samples for SEM were then mounted on stubs, allowed to off-gas in a vacuum desiccator for at least two hours and sputter coated with gold. Samples were examined with an Amray 1910 FE Scanning Electron Microscope and digitally imaged using Semicaps 2000 software. For TEM, ultra-thin sections were examined using a Philips CM100 electron microscope at 60 kV. Images were recorded digitally using a Hamamatsu ORCA-HR digital camera system operated with AMT software (Advanced Microscopy Techniques Corp., Danvers, MA).

### Differential proteomic analysis

Colonoids were isolated from Matrigel using 2mM EDTA for 15 minutes and then exposed to Radioimmunoprecipitation assay (RIPA) - lysis and extraction buffer (Pierce, # 89901; ThermoFisher Scientific) for protein isolation, as described in our previous reports (10,12). Proteomic experiments were carried out in the Proteomics Resource Facility (PRF) in the Department of Pathology at the University of Michigan, employing mass spectrometry-based Tandem Mass Tag (TMT, ThermoFisher Scientific). Fifty micrograms of colonoid protein from each condition (three subjects assessed separately) was digested separately with trypsin and individual samples labeled with one of 6 isobaric mass tags following the manufacturer’s protocol. After labeling, equal amounts of peptide from each condition were mixed together. In order to achieve in-depth characterization of the proteome, the labeled peptides were fractionated using 2D-LC (basic pH reverse-phase separation followed by acidic pH reverse phase) and analyzed on a high-resolution, tribrid mass spectrometer (Orbitrap Fusion Tribrid, ThermoFisher Scientific) using conditions optimized at the PRF. MultiNotch MS3 analysis (15) was employed to obtain accurate quantitation of the identified proteins/peptides. Data analysis was performed using Proteome Discoverer (v 2.3, ThermoFisher Scientific). MS2 spectra were searched against UniProt human protein database (20353 sequences; downloaded on 06/20/2019) using the following search parameters: MS1 and MS2 tolerance were set to 10 ppm and 0.6 Da, respectively; carbamidomethylation of cysteines (57.02146 Da) and TMT labeling of lysine and N-termini of peptides (229.16293 Da) were considered static modifications; oxidation of methionine (15.9949 Da) and deamidation of asparagine and glutamine (0.98401 Da) were considered variable. Identified proteins and peptides were filtered to retain only those that passed ≤2% false discovery rate (FDR) threshold of detection. Quantitation was performed using high-quality MS3 spectra (Average signal-to-noise ratio of 9 and <40% isolation interference). Differential protein expression between conditions, normalizing to control (0.25mM calcium) for each subject’s specimens separately was established using edgeR (16). Then, resulting data from the three data sets were combined. Proteins names were retrieved using Uniprot.org, and reactome V70 (reactome.org) was used for pathway enrichment analyses (17). Only Proteins with a ≤2% FDR confidence of detection were included in the analyses. The initial analysis was targeted towards differentiation, barrier-related, cell-cell and cell-matrix adhesion proteins. Follow-up analysis involved an unbiased proteome-wide screen of all proteins modified by intervention in relation to control.

### Cytokine assay

Approximately 400 μL of culture medium was collected at the time of harvest. A panel of pro-inflammatory cytokines, including tumor necrosis factor-α (TNF-α), interleukin-1β (IL-1β), IL-6, IL-8 and macrophage chemotactic peptide-1 (MCP-1) was assessed. Levels of each were determined using the Bio-Plex bead-based cytokine assay from Bio-Rad Laboratories (Hercules, CA).

### Statistical analysis

Group means and standard deviations were obtained for discrete morphological and immunohistochemical features as well as for individual protein values (proteomic analysis) and cytokine values. Data generated in this way were analyzed by analysis of variance (ANOVA) followed by unpaired t-test (two-tailed) for comparison using GraphPad Prism version 8. Proteome Discoverer (v 2.3) also computed an abundance ratio *p* value based on a nested design of biological replicates. It presents the *p* value of the sample group calculated by running the Tukey HSD test (post hoc) after an ANOVA test. Pathways enrichment data reflect Reactome-generated *p*-values based on the number of entities identified in a given pathway as compared to total proteins responsible for that pathway. Data were considered significant at *p*<0.05.

## ETHICAL CONSIDERATIONS

The submitted manuscript is an original contribution and has not been previously published. It is not under consideration for publication elsewhere. The current manuscript has been deposited to a preprint server “medRxiv” prior to this submission which is consistent with the journal’s policy. All authors have contributed towards this project to claim authorship according to ICMJE criteria. The authors have no financial or personal conflict of interest to declare.

For the current study, de-identified colon tissue was collected during surveillance colonoscopy. Tissue collection was done under a protocol approved by the Institutional Review Board at the University of Michigan (IRBMED). Patients’ protected health information was not shared with the investigators. Each subject signed a written informed consent document prior to the procedure allowing the use of the colonic tissue for research purposes.

## RESULTS

### Establishment of colonoid cultures from UC biopsies

Colonic tissue (duplicate 2-mm biopsies) was obtained by endoscopy from the sigmoid colon of 11 subjects with a diagnosis of UC. Five of the subjects had active disease at time of biopsy. The other six participants were considered to be in remission; though all had disease-related symptoms (or active flare) within the past four years of tissue collection. Relevant demographic and clinical information is presented in Table 1 Tissue was established in colonoid culture as described in the Materials and Methods Section. Two of the active-lesion samples failed to grow and were eventually discarded. Successful cultures were established from the remaining nine tissue samples. The “in-remission” colonic tissue samples were satisfactorily-expanded and were ready for experimentation after two to three weeks – similar to tissue from healthy individuals (10). Tissue from “active lesion” samples required 3-5 weeks to reach this stage.

**Table 1.**
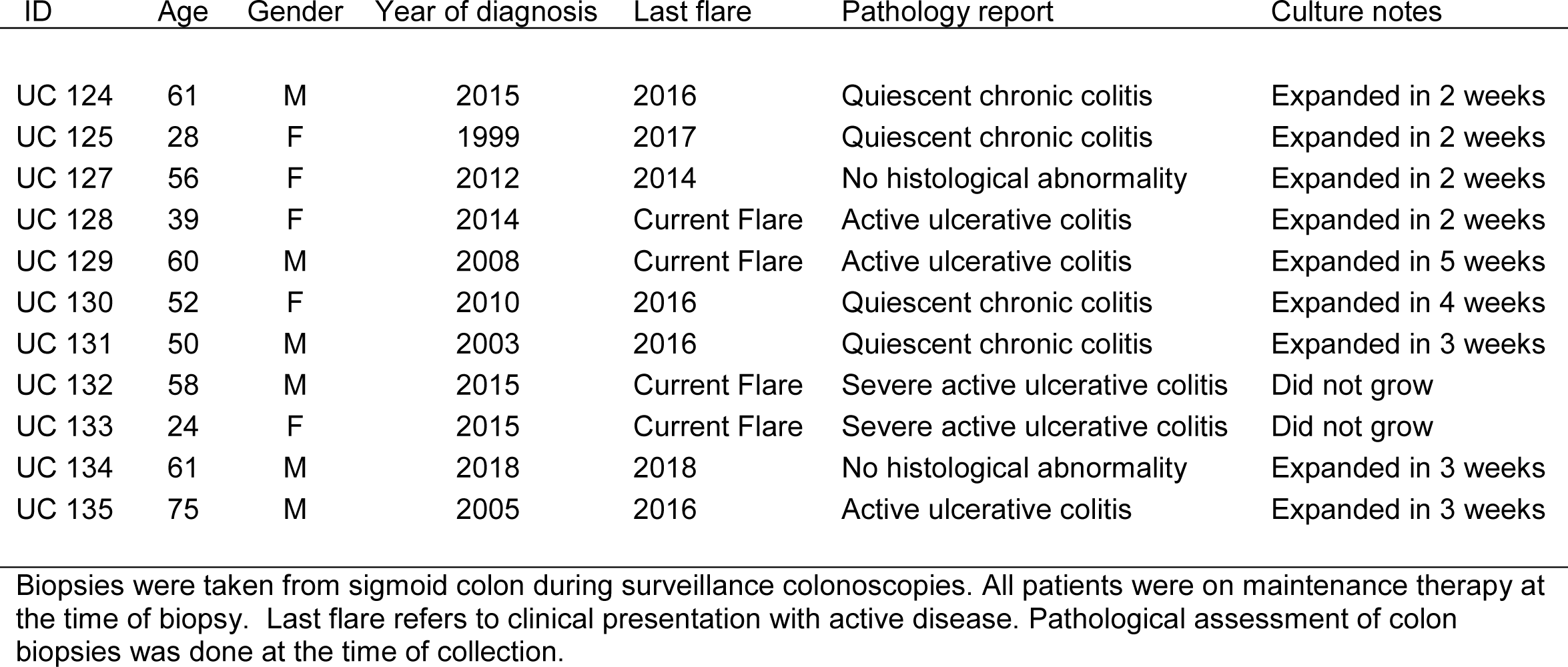
Demographic information, disease status, pathology report and culture notes.

| ID | Age | Gender | Year of diagnosis | Last flare | Pathology report | Culture notes |
| --- | --- | --- | --- | --- | --- | --- |
| UC 124 | 61 | M | 2015 | 2016 | Quiescent chronic colitis | Expanded in 2 weeks |
| UC 125 | 28 | F | 1999 | 2017 | Quiescent chronic colitis | Expanded in 2 weeks |
| UC 127 | 56 | F | 2012 | 2014 | No histological abnormality | Expanded in 2 weeks |
| UC 128 | 39 | F | 2014 | Current Flare | Active ulcerative colitis | Expanded in 2 weeks |
| UC 129 | 60 | M | 2008 | Current Flare | Active ulcerative colitis | Expanded in 5 weeks |
| UC 130 | 52 | F | 2010 | 2016 | Quiescent chronic colitis | Expanded in 4 weeks |
| UC 131 | 50 | M | 2003 | 2016 | Quiescent chronic colitis | Expanded in 3 weeks |
| UC 132 | 58 | M | 2015 | Current Flare | Severe active ulcerative colitis | Did not grow |
| UC 133 | 24 | F | 2015 | Current Flare | Severe active ulcerative colitis | Did not grow |
| UC 134 | 61 | M | 2018 | 2018 | No histological abnormality | Expanded in 3 weeks |
| UC 135 | 75 | M | 2005 | 2016 | Active ulcerative colitis | Expanded in 3 weeks |

Biopsies were taken from sigmoid colon during surveillance colonoscopies. All patients were on maintenance therapy at the time of biopsy. Last flare refers to clinical presentation with active disease. Pathological assessment of colon biopsies was done at the time of collection.

### Effects of mineral intervention on gross and microscopic features

Once established, colonoids were incubated for a two-week period (with subculture at the end of week-one) under control conditions (0.25 mM calcium) or in the presence of the multi-mineral intervention formulated to provide 1.5 – 4.5 mM calcium. Phase-contrast images and SEM images taken at harvest (Figure 1a-h) identified no gross morphological changes attributable to the intervention. In both control and treated cultures, individual colonoids appeared as round or egg-shaped structures. Most colonoids consisted of a single structure but some were multi-lobed. In most places, the colonoids still had a layer of the Matrigel substrate attached, but in other places, the substrate was gone and individual colonoid cells could be seen. At the beginning of the treatment phase, and immediately after the first subculture, colonoid structures were approximately 40 μm in size (on average). Over the culture period, they increased to approximately 400 μm in diameter. Overall, UC-derived colonoid appearance and growth characteristics were similar to what has been reported previously with colonoids established from tissue of healthy individuals (10).

**Figure 1.**
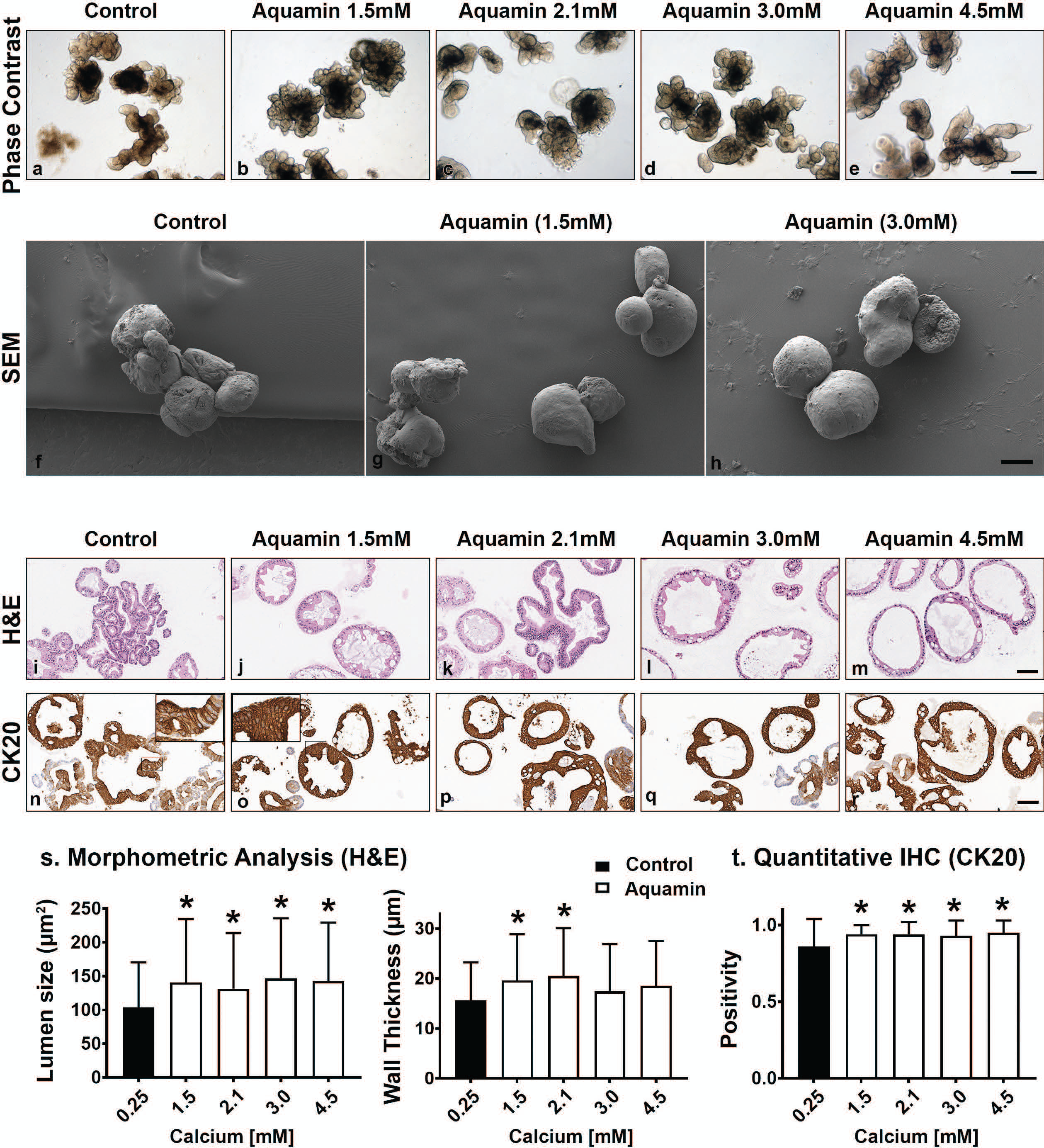
**UC colonoid appearance. Phase-contrast microscopy (a-e):** At the end of the incubation period, intact colonoids were examined by phase-contrast microscopy. Colonoids were present as thick-walled structures with few surface buds. A wide range of sizes and shapes were seen under all conditions. Bar=400µm. **Scanning electron microscopy (f-h)**: Scanning electron microscopy confirmed the presence of smooth surface and few buds in colonoids maintained under low-calcium conditions (f). Aquamin^®^-treated colonoids (g,h) were similar to those maintained in the low-calcium medium. Bar =100 μm. **Histological features (i-m)**: At the end of the incubation period, colonoids were examined by light microscopy after staining with hematoxylin and eosin. Under low-calcium conditions (i), the colonoids were found to be crypts of varying size with a single layer of epithelial cells surrounding a central lumen. Tiny crypts (with as few as 20 cells in cross section) were seen. In the presence of Aquamin^®^ (j-m), larger crypts made up of columnar epithelial cells surrounding a large, often irregular-shaped lumen were seen. Goblet cells were apparent. Bar=100μm. **CK20 expression (n-r)**: Immunohistology revealed high-expression of CK20 under all conditions. Bar=100µm. **Quantification of morphological features and CK20 expression (s and t)**: Lumen size and wall thickness (s): Means and standard deviations are based on n=3 subjects and 87-143 individual crypts per condition. Asterisks (*) indicate statistical significance from control at p<0.05. CK20 expression (t): Means and standard deviations are based on n=3 subjects and 81-120 individual crypts per condition. Asterisks (*) indicate statistical significance from control at p<0.05.

Although there were no apparent differences in gross morphology, differences in differentiation features were revealed by quantitative histology and immunohistology. As seen in hematoxylin and eosin-stained tissue sections (Figure 1i-m), colonoids maintained for two weeks in the presence of the multi-mineral intervention had a larger lumen diameter and greater wall thickness than control. This was confirmed morphometrically (Figure 1s). Expression of the differentiation marker, CK20 (Figure 1n-r), was substantial under control conditions, but still increased (albeit, modestly) in response to treatment (Figure 1t). These findings from the gross and microscopic analyses allow us to conclude that colonoids established from UC tissue reveal a high degree of differentiation in the control medium. Even with the high background, exposure to the combination of minerals in Aquamin^®^ “pushes” differentiation further, which is evident by the change in morphology and increase in the expression of CK20.

Epithelial differentiation is linked with growth reduction (18). As part of our evaluation, we assessed Ki67 expression in control and treated colonoids. A decrease in Ki67 expression was seen with treatment (Supplement Figure 1a-e, k). Levels under both control and treated conditions were comparable to what was seen previously in colonoids from healthy subjects (10), but lower than what we observed with colon adenomas in culture (12).

### Effects of Aquamin^®^ on proteins involved in differentiation, cell-cell / cell-matrix adhesion and barrier formation

A Tandem Mass Tag mass spectrometry-based approach was used to assess the effects of Aquamin^®^ on proteins involved in differentiation and barrier formation. Table 2 provides a list of proteins that are part of the differentiation process and / or contribute to the barrier. There are several things to note. First, CK20 up-regulation was seen with Aquamin^®^-treatment; this is consistent with the immunohistochemical expression data shown as part of Figure 1. A number of other differentiation-related proteins were also up-regulated in Aquamin^®^-treated colonoids (Table 2), but only with CK80 was the level of up-regulation comparable to that of CK20. Several additional keratins to those shown in Table 2 were also detected but did not change significantly with treatment or actually decreased with intervention.

Second, several proteins that make up tight junctions were detected but overall showed little change in response to mineral-intervention. Claudin-4 was the most responsive tight junction protein but was only increased by 1.4-fold at maximum. In contrast, desmosomal proteins were strongly up-regulated with increasing concentrations of the multi-mineral product. Several cadherin family members (adherens junction proteins) were also up-regulated. The major exception was cadherin-1 (E-cadherin), a protein closely associated with tight junction organization and that is known to have low sensitivity to changes in calcium concentration (19).

Immunohistology was used to confirm desmoglein-2 expression and change with intervention. As expected based on proteomic analysis, there was little expression under control conditions but strong up-regulation with intervention (Figure 2a-e, i). Control and Aquamin^®^-treated colonoid samples were also evaluated by TEM (Figure 2f-h). Desmosomes were visible under all conditions, but desmosome numbers increased measurably in treated colonoids as compared to control (white arrows) (Figure 2f-h, j). Desmosomes from the treated samples also tended to be wider and more electron-dense. As part of the immunohistological evaluation, cadherin-17 expression was evaluated as an adherens junction protein. Cadherin-17 was readily detected under low-calcium conditions in colonoid sections, but even with the high background, an increase with intervention was seen (Supplement Figure 1f-j)

**Figure 2.**
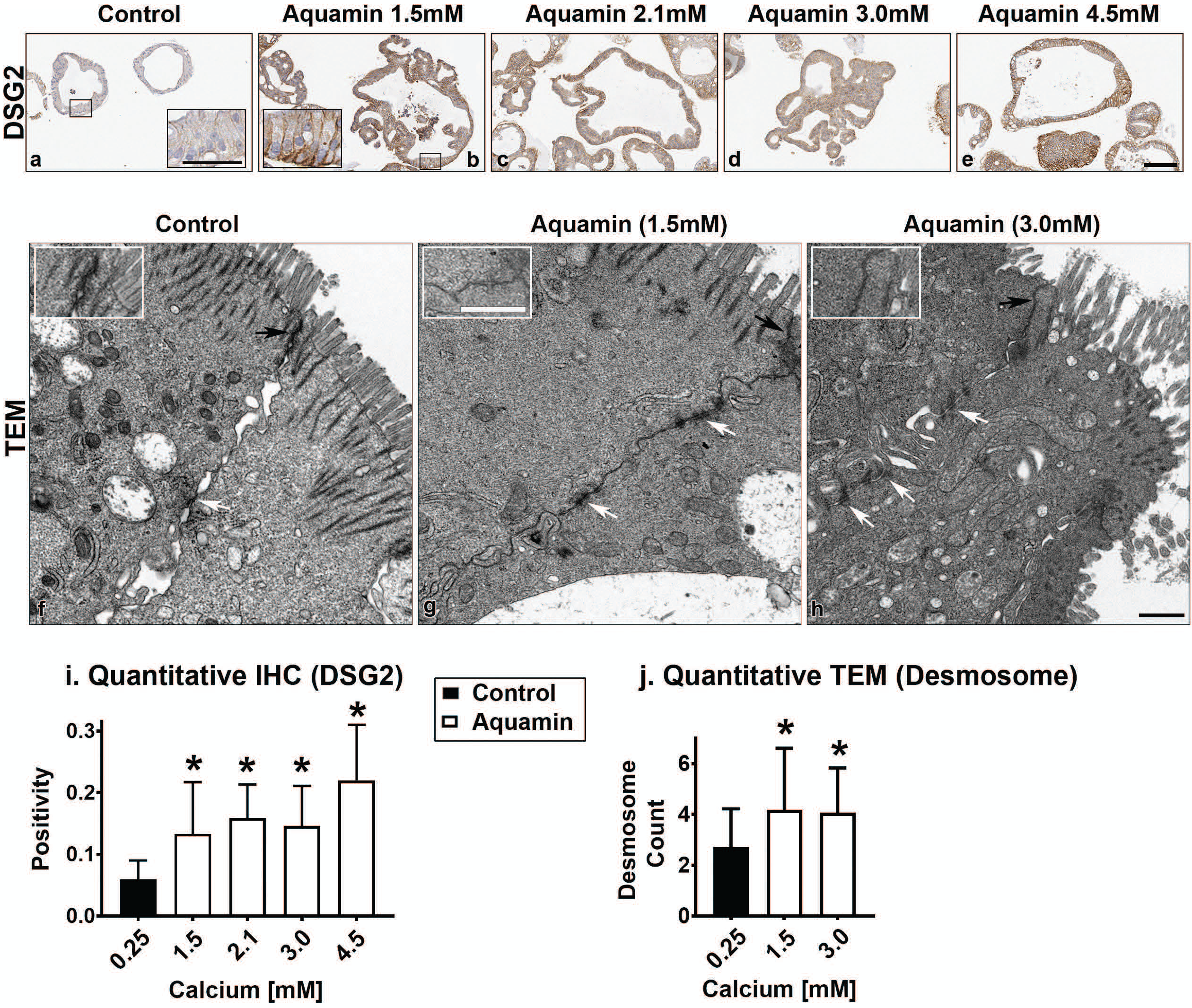
**Desmoglein-2 and desmosomes. Immunohistology (a-e):** At the end of the incubation period, tissue sections were stained for desmoglein-2 and examined. Staining was diffuse and intracellular in colonoids maintained under low-calcium conditions. Staining was more intense in sections from Aquamin^®^-treated colonoids. Staining was prominent along the basolateral border in treated colonoids as seen in the inset. Bar=200 μm, inset bar=50 μm. **Transmission electron microscopy (f-h)**: At the end of the incubation period, ultra-thin sections were examined for desmosomes and other cell surface structures. Desmosomes were present in all conditions (white arrows) but a higher density of desmosomes along the lateral surface (cellular junctions between two cells) could be seen with intervention. Under all conditions, tight junctions were evident on the luminal surface (black arrows and insets). Magnification: 5,000X; Bars=600 nm. **Quantification of desmoglein-2 expression (i) and desmosome counts (j)**: Immunostaining (i) results are means and standard deviations based on n=3 subjects and 85-139 individual crypts per condition. Asterisks (*) indicate statistical significance from control at p<0.05. The desmosome count was conducted at 5000X (n=3 subjects with 7-18 images per subject) to obtain the actual number (means and SD) of desmosomes present in each high-power section. Asterisks indicate statistical significance from control at p<0.05 level.

In addition to proteins that constitute the major cell-cell adhesion complexes in the colonic epithelium, several additional proteins that contribute to cellular adhesive functions and to the structural barrier were also found to be up-regulated in the mineral-treated colonoids (Table 2). These include proteins making up the basement membrane (laminin α, β and γ chains, nidogen-1 [entactin] and perlecan [basement membrane-specific heparin sulfate proteoglycan]) as well as a protein that contributes to hemidesmosome formation and basement membrane attachment (e.g., plectin).

Proteins found at the apical surface – mucins and trefoils – are also shown in Table 2. Certain of these (e.g., mucin-5B & -13 as well as trefoil-1 & -2) were significantly elevated in response to treatment. Trefoils are proteins that interact with mucins to stabilize the mucous layer (20,21). Finally, a number of carcinoembryonic antigen related – cell adhesion molecules (CEACAMs) were strongly up-regulated in response to Aquamin^®^. This family of proteins demonstrated more subject-to-subject variability than was seen with most of the other moieties. In addition, whereas most of the relevant proteins increased with increasing mineral concentration, several CEACAMs showed a peak response to Aquamin^®^ at a level providing 2.1 or 3.0 mM calcium and fell off at 4.5 mM.

**Table 2A.**
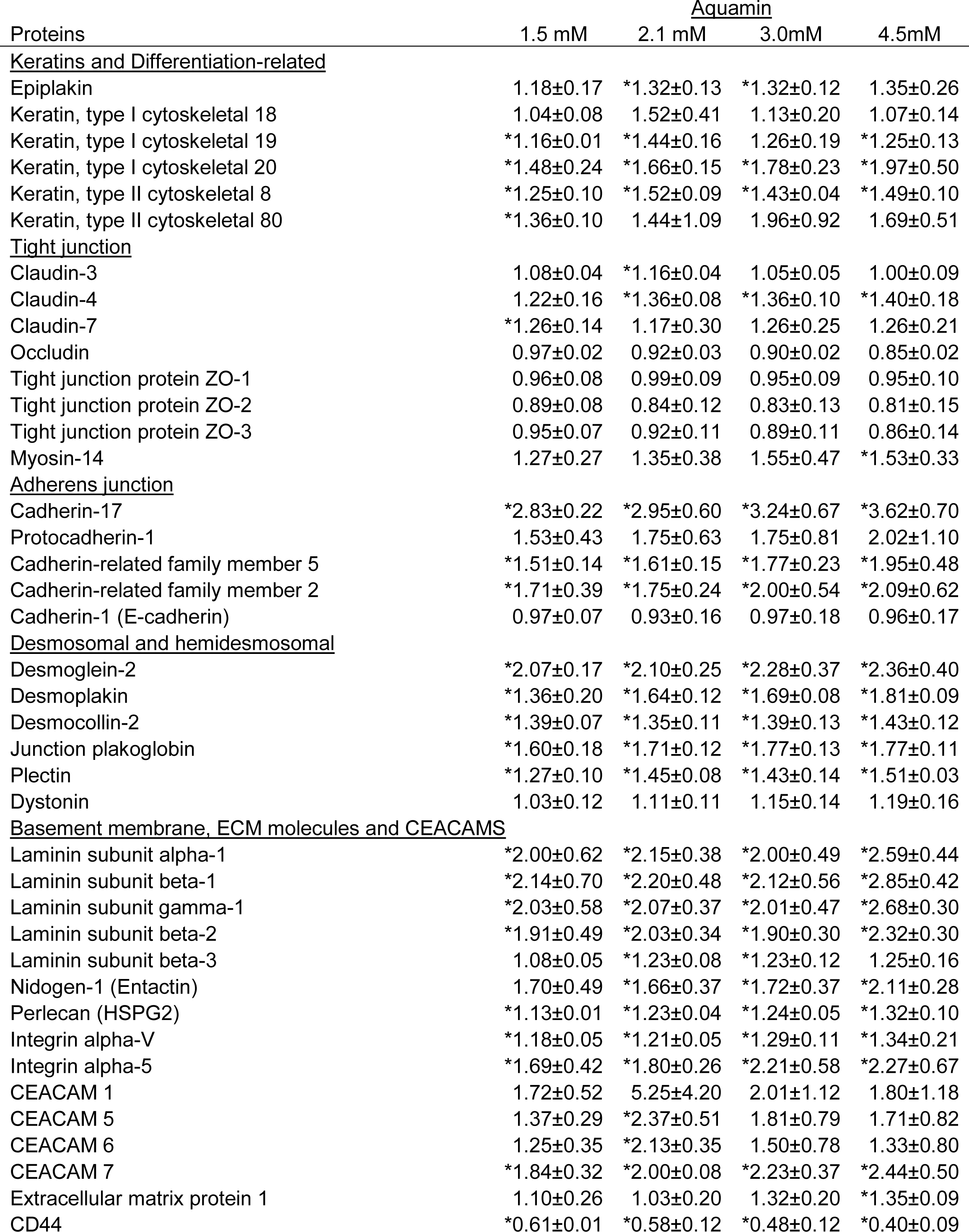
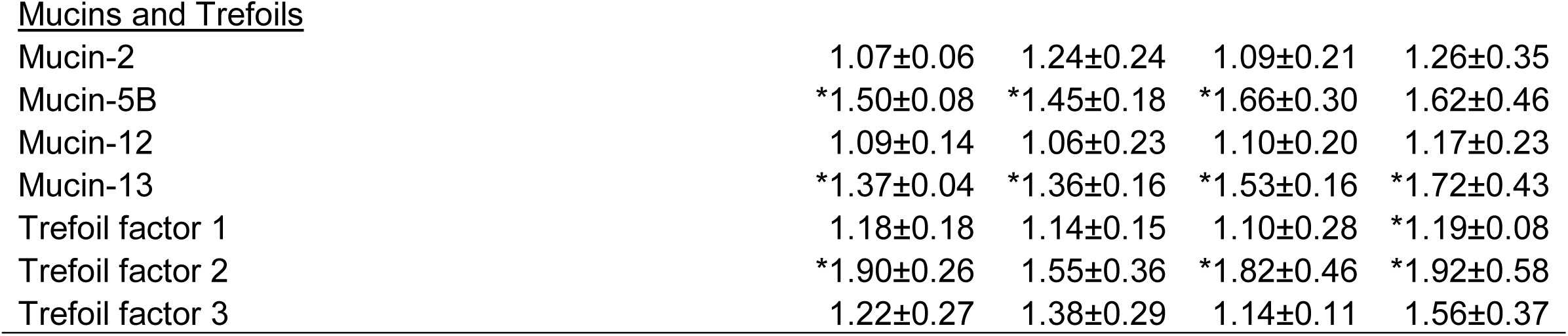
Proteins involved in differentiation and barrier formation.

| Proteins | Aquamin |  |  |  |
| --- | --- | --- | --- | --- |
|  | 1.5 mM | 2.1 mM | 3.0mM | 4.5mM |
| <u>Keratins and Differentiation-related</u> |  |  |  |  |
| Epiplakin | 1.18±0.17 | *1.32±0.13 | *1.32±0.12 | 1.35±0.26 |
| Keratin, type I cytoskeletal 18 | 1.04±0.08 | 1.52±0.41 | 1.13±0.20 | 1.07±0.14 |
| Keratin, type I cytoskeletal 19 | *1.16±0.01 | *1.44±0.16 | 1.26±0.19 | *1.25±0.13 |
| Keratin, type I cytoskeletal 20 | *1.48±0.24 | *1.66±0.15 | *1.78±0.23 | *1.97±0.50 |
| Keratin, type II cytoskeletal 8 | *1.25±0.10 | *1.52±0.09 | *1.43±0.04 | *1.49±0.10 |
| Keratin, type II cytoskeletal 80 | *1.36±0.10 | 1.44±1.09 | 1.96±0.92 | 1.69±0.51 |
| <u>Tight junction</u> |  |  |  |  |
| Claudin-3 | 1.08±0.04 | *1.16±0.04 | 1.05±0.05 | 1.00±0.09 |
| Claudin-4 | 1.22±0.16 | *1.36±0.08 | *1.36±0.10 | *1.40±0.18 |
| Claudin-7 | *1.26±0.14 | 1.17±0.30 | 1.26±0.25 | 1.26±0.21 |
| Occludin | 0.97±0.02 | 0.92±0.03 | 0.90±0.02 | 0.85±0.02 |
| Tight junction protein ZO-1 | 0.96±0.08 | 0.99±0.09 | 0.95±0.09 | 0.95±0.10 |
| Tight junction protein ZO-2 | 0.89±0.08 | 0.84±0.12 | 0.83±0.13 | 0.81±0.15 |
| Tight junction protein ZO-3 | 0.95±0.07 | 0.92±0.11 | 0.89±0.11 | 0.86±0.14 |
| Myosin-14 | 1.27±0.27 | 1.35±0.38 | 1.55±0.47 | *1.53±0.33 |
| <u>Adherens junction</u> |  |  |  |  |
| Cadherin-17 | *2.83±0.22 | *2.95±0.60 | *3.24±0.67 | *3.62±0.70 |
| Protocadherin-1 | 1.53±0.43 | 1.75±0.63 | 1.75±0.81 | 2.02±1.10 |
| Cadherin-related family member 5 | *1.51±0.14 | *1.61±0.15 | *1.77±0.23 | *1.95±0.48 |
| Cadherin-related family member 2 | *1.71±0.39 | *1.75±0.24 | *2.00±0.54 | *2.09±0.62 |
| Cadherin-1 (E-cadherin) | 0.97±0.07 | 0.93±0.16 | 0.97±0.18 | 0.96±0.17 |
| <u>Desmosomal and hemidesmosomal</u> |  |  |  |  |
| Desmoglein-2 | *2.07±0.17 | *2.10±0.25 | *2.28±0.37 | *2.36±0.40 |
| Desmoplakin | *1.36±0.20 | *1.64±0.12 | *1.69±0.08 | *1.81±0.09 |
| Desmocollin-2 | *1.39±0.07 | *1.35±0.11 | *1.39±0.13 | *1.43±0.12 |
| Junction plakoglobin | *1.60±0.18 | *1.71±0.12 | *1.77±0.13 | *1.77±0.11 |
| Plectin | *1.27±0.10 | *1.45±0.08 | *1.43±0.14 | *1.51±0.03 |
| Dystonin | 1.03±0.12 | 1.11±0.11 | 1.15±0.14 | 1.19±0.16 |
| <u>Basement membrane, ECM molecules and CEACAMS</u> |  |  |  |  |
| Laminin subunit alpha-1 | *2.00±0.62 | *2.15±0.38 | *2.00±0.49 | *2.59±0.44 |
| Laminin subunit beta-1 | *2.14±0.70 | *2.20±0.48 | *2.12±0.56 | *2.85±0.42 |
| Laminin subunit gamma-1 | *2.03±0.58 | *2.07±0.37 | *2.01±0.47 | *2.68±0.30 |
| Laminin subunit beta-2 | *1.91±0.49 | *2.03±0.34 | *1.90±0.30 | *2.32±0.30 |
| Laminin subunit beta-3 | 1.08±0.05 | *1.23±0.08 | *1.23±0.12 | 1.25±0.16 |
| Nidogen-1 (Entactin) | 1.70±0.49 | *1.66±0.37 | *1.72±0.37 | *2.11±0.28 |
| Perlecan (HSPG2) | *1.13±0.01 | *1.23±0.04 | *1.24±0.05 | *1.32±0.10 |
| Integrin alpha-V | *1.18±0.05 | *1.21±0.05 | *1.29±0.11 | *1.34±0.21 |
| Integrin alpha-5 | *1.69±0.42 | *1.80±0.26 | *2.21±0.58 | *2.27±0.67 |
| CEACAM 1 | 1.72±0.52 | 5.25±4.20 | 2.01±1.12 | 1.80±1.18 |
| CEACAM 5 | 1.37±0.29 | *2.37±0.51 | 1.81±0.79 | 1.71±0.82 |
| CEACAM 6 | 1.25±0.35 | *2.13±0.35 | 1.50±0.78 | 1.33±0.80 |
| CEACAM 7 | *1.84±0.32 | *2.00±0.08 | *2.23±0.37 | *2.44±0.50 |
| Extracellular matrix protein 1 | 1.10±0.26 | 1.03±0.20 | 1.32±0.20 | *1.35±0.09 |
| CD44 | *0.61±0.01 | *0.58±0.12 | *0.48±0.12 | *0.40±0.09 |
| Mucins and Trefoils |  |  |  |  |
| Mucin-2 | 1.07±0.06 | 1.24±0.24 | 1.09±0.21 | 1.26±0.35 |
| Mucin-5B | *1.50±0.08 | *1.45±0.18 | *1.66±0.30 | 1.62±0.46 |
| Mucin-12 | 1.09±0.14 | 1.06±0.23 | 1.10±0.20 | 1.17±0.23 |
| Mucin-13 | *1.37±0.04 | *1.36±0.16 | *1.53±0.16 | *1.72±0.43 |
| Trefoil factor 1 | 1.18±0.18 | 1.14±0.15 | 1.10±0.28 | *1.19±0.08 |
| Trefoil factor 2 | *1.90±0.26 | 1.55±0.36 | *1.82±0.46 | *1.92±0.58 |
| Trefoil factor 3 | 1.22±0.27 | 1.38±0.29 | 1.14±0.11 | 1.56±0.37 |

**Table 2B.**
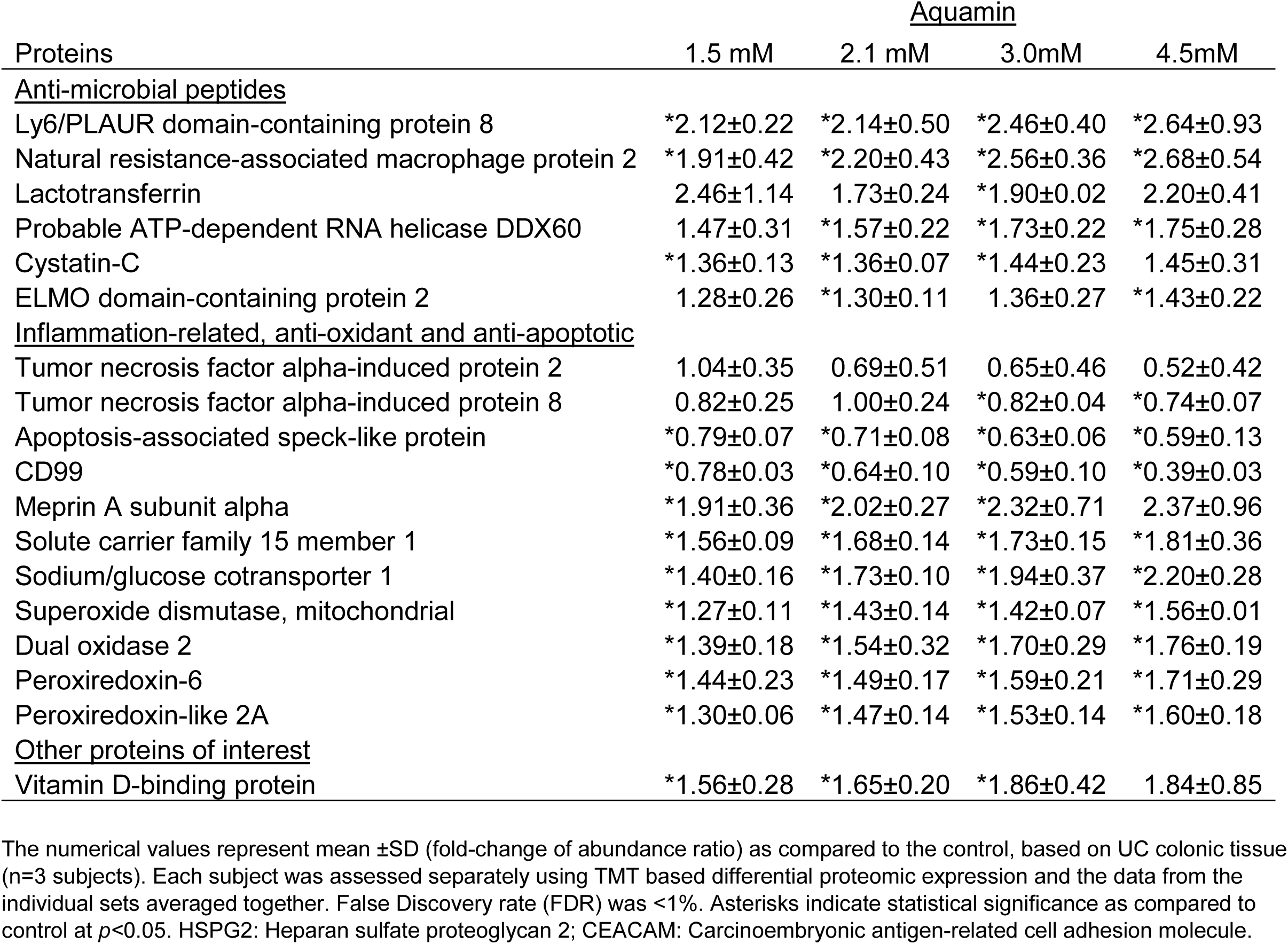
Inflammation-related proteins.

| Proteins | <u>Aquamin</u> |  |  |  |
| --- | --- | --- | --- | --- |
|  | 1.5 mM | 2.1 mM | 3.0mM | 4.5mM |
| <u>Anti-microbial peptides</u> |  |  |  |  |
| Ly6/PLAUR domain-containing protein 8 | *2.12±0.22 | *2.14±0.50 | *2.46±0.40 | *2.64±0.93 |
| Natural resistance-associated macrophage protein 2 | *1.91±0.42 | *2.20±0.43 | *2.56±0.36 | *2.68±0.54 |
| Lactotransferrin | 2.46±1.14 | 1.73±0.24 | *1.90±0.02 | 2.20±0.41 |
| Probable ATP-dependent RNA helicase DDX60 | 1.47±0.31 | *1.57±0.22 | *1.73±0.22 | *1.75±0.28 |
| Cystatin-C | *1.36±0.13 | *1.36±0.07 | *1.44±0.23 | 1.45±0.31 |
| ELMO domain-containing protein 2 | 1.28±0.26 | *1.30±0.11 | 1.36±0.27 | *1.43±0.22 |
| <u>Inflammation-related, anti-oxidant and anti-apoptotic</u> |  |  |  |  |
| Tumor necrosis factor alpha-induced protein 2 | 1.04±0.35 | 0.69±0.51 | 0.65±0.46 | 0.52±0.42 |
| Tumor necrosis factor alpha-induced protein 8 | 0.82±0.25 | 1.00±0.24 | *0.82±0.04 | *0.74±0.07 |
| Apoptosis-associated speck-like protein | *0.79±0.07 | *0.71±0.08 | *0.63±0.06 | *0.59±0.13 |
| CD99 | *0.78±0.03 | *0.64±0.10 | *0.59±0.10 | *0.39±0.03 |
| Meprin A subunit alpha | *1.91±0.36 | *2.02±0.27 | *2.32±0.71 | 2.37±0.96 |
| Solute carrier family 15 member 1 | *1.56±0.09 | *1.68±0.14 | *1.73±0.15 | *1.81±0.36 |
| Sodium/glucose cotransporter 1 | *1.40±0.16 | *1.73±0.10 | *1.94±0.37 | *2.20±0.28 |
| Superoxide dismutase, mitochondrial | *1.27±0.11 | *1.43±0.14 | *1.42±0.07 | *1.56±0.01 |
| Dual oxidase 2 | *1.39±0.18 | *1.54±0.32 | *1.70±0.29 | *1.76±0.19 |
| Peroxiredoxin-6 | *1.44±0.23 | *1.49±0.17 | *1.59±0.21 | *1.71±0.29 |
| Peroxiredoxin-like 2A | *1.30±0.06 | *1.47±0.14 | *1.53±0.14 | *1.60±0.18 |
| <u>Other proteins of interest</u> |  |  |  |  |
| Vitamin D-binding protein | *1.56±0.28 | *1.65±0.20 | *1.86±0.42 | 1.84±0.85 |

The numerical values represent mean ±SD (fold-change of abundance ratio) as compared to the control, based on UC colonic tissue (n=3 subjects). Each subject was assessed separately using TMT based differential proteomic expression and the data from the individual sets averaged together. False Discovery rate (FDR) was <1%. Asterisks indicate statistical significance as compared to control at $p < 0.05$ . HSPG2: Heparan sulfate proteoglycan 2; CEACAM: Carcinoembryonic antigen-related cell adhesion molecule.

### Effects of Aquamin^®^ on proteins with anti-microbial activity and proteins related to inflammation

In addition to the structural proteins described above, the proteomic analysis also identified up-regulation of several proteins with known anti-microbial properties and modulation of additional proteins involved in the inflammatory response. Proteins with pro-inflammatory effects were decreased while proteins with anti-inflammatory, anti-oxidant and anti-apoptotic activity were uniformly increased (Table 2). Among the list of up-regulated proteins were meprin A, a metalloproteinase whose presence in colon tissue is reduced in UC, a member of the solute carrier family-15 family and a protein known as sodium/glucose cotransporter-1 (SGLT-1). All of these proteins are thought to have an anti-inflammatory role in colitis. Finally, the major vitamin D binding protein was also strongly increased with intervention. Vitamin D signaling is important to calcium uptake and utilization. Supplement Text File 1 provide a list of citations to justify inclusion of the anti-microbial and inflammation-modulating proteins in Table 2.

### Unbiased assessment of proteins up-regulated or down-regulated with multi-mineral Intervention

The proteins presented in Table 2 were chosen based on a known or suspected role in differentiation and barrier formation or in consideration of their relevance to the pathophysiology of inflammatory bowel disease. In parallel, we conducted an unbiased search of the data base for all proteins up- or down-regulated by Aquamin^®^ treatment. The list of proteins that met the criteria of 1.8-fold increase or decrease across the three data sets (i.e., from each subject) with a ≤ 2% FDR are shown in Tables 3 and 4 along with pathways linked to these proteins. Twenty-nine proteins were up-regulated in all three specimens (Table 3). Of interest, the majority of these were proteins involved in differentiation and barrier formation – i.e., the same proteins presented in Table 2. There were 50 down-regulated proteins that met this criterion. As can be seen from Table 4, many of these were involved in nucleotide metabolism, proliferation-related signaling and proliferation. Supplement Figure 2 shows the number of up- and down-regulated proteins across the three data sets at each of four levels of Aquamin^®^. The Venn plots in the upper panel show overlap at each Aquamin^®^ level separately among three subjects, and the plot in the lower level merges common proteins at all Aquamin^®^ concentrations and the three data sets. Supplement Table 3 identifies individual proteins common at different levels of Aquamin^®^. Lastly, Supplement Table 4 presents a comprehensive list of proteins (308 proteins in all and up- or down-regulated) that met the criterion of being significant with a *p* value of 0.05 or less across all conditions of Aquamin^®^ as compared to control (regardless of fold-change).

**Table 3A.** Up-regulated proteins: Unbiased proteomic screen.

| Proteins | Aquamin |  |  |  |
| --- | --- | --- | --- | --- |
|  | 1.5 mM | 2.1 mM | 3.0mM | 4.5mM |
| Nuclear receptor 0B1<sup>#</sup> | 2.59±1.13 | 5.71±3.91 | 4.69±2.45 | 6.68±4.11 |
| CEACAM 1 | 1.72±0.52 | 5.25±4.20 | 2.01±1.12 | 1.80±1.18 |
| Alpha-2-HS-glycoprotein | 2.24±0.87 | 2.39±1.18 | 2.59±1.69 | 2.48±0.95 |
| Cadherin-17 | *2.83±0.22 | *2.95±0.60 | *3.23±0.67 | *3.62±0.70 |
| Calcium-activated chloride channel regulator 4 | *2.03±0.28 | *2.24±0.51 | *2.31±0.27 | *3.01±0.74 |
| Zinc transporter ZIP4 | *2.01±0.55 | *2.08±0.46 | *2.63±0.77 | *2.83±0.96 |
| Ly6/PLAUR domain-containing protein 8 | *2.12±0.22 | *2.14±0.50 | *2.46±0.40 | *2.64±0.93 |
| Meprin A subunit alpha | *1.91±0.36 | *2.02±0.27 | *2.32±0.71 | 2.37±0.96 |
| Natural resistance-associated macrophage protein 2 | *1.91±0.42 | *2.20±0.43 | *2.56±0.36 | *2.68±0.54 |
| Laminin subunit beta-1 | *2.14±0.70 | *2.20±0.48 | *2.12±0.56 | *2.85±0.42 |
| Laminin subunit alpha-1 | *2.00±0.62 | *2.15±0.38 | *2.00±0.49 | *2.59±0.44 |
| Integrin alpha-5 | *1.69±0.42 | *1.80±0.26 | *2.21±0.58 | *2.27±0.67 |
| CEACAM 7 | *1.83±0.32 | *2.00±0.08 | *2.23±0.37 | *2.44±0.50 |
| Apolipoprotein A-I | 1.62±0.61 | *1.96±0.46 | *2.39±0.28 | *2.68±0.30 |
| CEACAM 5 | 1.37±0.29 | *2.37±0.50 | 1.81±0.79 | 1.71±0.82 |
| Laminin subunit gamma-1 | *2.03±0.58 | *2.07±0.37 | *2.01±0.47 | *2.68±0.30 |
| Desmoglein-2 | *2.07±0.17 | *2.10±0.25 | *2.28±0.37 | *2.36±0.40 |
| Fucose mutarotase | *1.44±0.13 | *1.76±0.25 | *1.83±0.19 | *2.04±0.58 |
| Laminin subunit beta-2 | *1.91±0.49 | *2.03±0.34 | *1.90±0.30 | *2.32±0.30 |
| Cyclic AMP-dependent transcription factor ATF-7<sup>#</sup> | *1.37±0.13 | *2.25±0.37 | *1.76±0.45 | 1.74±0.57 |
| CEACAM 6 | 1.25±0.35 | *2.13±0.35 | 1.50±0.78 | 1.33±0.80 |
| Sodium/glucose cotransporter 1 | *1.40±0.16 | *1.73±0.10 | *1.94±0.37 | *2.20±0.28 |
| Nidogen-1 | 1.70±0.49 | *1.66±0.37 | *1.72±0.37 | *2.11±0.28 |
| Copper-transporting ATPase 2 | 1.39±0.52 | *1.62±0.22 | *1.94±0.16 | *2.28±0.04 |
| ENPP3 | *1.41±0.15 | *1.60±0.18 | *1.81±0.19 | *2.04±0.25 |
| Aminopeptidase N | *1.91±0.21 | *2.01±0.25 | *1.85±0.27 | *2.00±0.36 |
| Beta-glucuronidase | *1.29±0.06 | *1.51±0.23 | *1.51±0.17 | *2.00±0.27 |
| Hydroxymethylglutaryl-CoA synthase, mitochondrial | *1.52±0.12 | *1.74±0.23 | *1.95±0.22 | *2.09±0.18 |
| Inter-alpha-trypsin inhibitor heavy chain H4 | *1.46±0.08 | *1.78±0.31 | *1.79±0.32 | *1.98±0.20 |

The numerical values represent mean ±SD (fold-change of abundance ratio) as compared to the control, based on UC colonic tissue (n=3 subjects). Each subject was assessed separately using TMT based differential proteomic expression and the data from the individual sets averaged together. False Discovery rate (FDR) was <1% except two proteins<sup>#</sup> with FDR<2%. Values were ordered from maximum to minimum fold-change based on the maximum response with any of the Aquamin® concentrations. To be included, values were up-regulated by at least 1.8-fold with at least one concentration of Aquamin® as compared to control and common in all three subjects. Asterisks indicate statistical significance as compared to control at $p<0.05$ . CEACAM: Carcinoembryonic antigen-related cell adhesion molecule; ENPP3: Ectonucleotide pyrophosphatase/phosphodiesterase family member 3.

**Table 3B.** Pathways significantly activated based on up-regulated Proteins.

| <u>Pathway name</u> | <u>p Value</u> | <u>FDR</u> |
| --- | --- | --- |
| Laminin interactions | 0.00000002 | 0.000002 |
| Fibronectin matrix formation | 0.000001 | 0.00004 |
| Extracellular matrix organization | 0.000001 | 0.00004 |
| MET activates PTK2 signaling | 0.000001 | 0.00004 |
| MET promotes cell motility | 0.000005 | 0.0001 |
| Post-translational protein phosphorylation | 0.00001 | 0.0002 |
| Regulation of IGF transport and uptake by IGF Binding Proteins (IGFBPs) | 0.00002 | 0.0003 |
| Non-integrin membrane-ECM interactions | 0.00002 | 0.0003 |
| ECM proteoglycans | 0.00005 | 0.0007 |
| Signaling by MET | 0.00006 | 0.0007 |
| L1CAM interactions | 0.0003 | 0.003 |
| Disorders of transmembrane transporters | 0.001 | 0.01 |
| Post-translational modification: synthesis of GPI-anchored proteins | 0.002 | 0.01 |
| Metal ion SLC transporters | 0.002 | 0.01 |
| MPS VII - Sly syndrome | 0.003 | 0.01 |
| Defective SLC39A4 causes acrodermatitis enteropathica, zinc-deficiency | 0.003 | 0.01 |
| Defective SLC11A2 causes hypochromic microcytic anemia | 0.003 | 0.01 |
| Defective SLC5A1 causes congenital glucose/galactose malabsorption | 0.003 | 0.01 |
| SLC transporter disorders | 0.003 | 0.01 |
| Cell surface interactions at the vascular wall | 0.004 | 0.02 |
| Defective ABCA1 causes Tangier disease | 0.005 | 0.02 |
| Response to elevated platelet cytosolic Ca<sup>2+</sup> | 0.005 | 0.02 |
| Degradation of the extracellular matrix | 0.006 | 0.02 |
| Neutrophil degranulation | 0.008 | 0.03 |
| Post-translational protein modification | 0.01 | 0.04 |
| Intestinal hexose absorption | 0.01 | 0.04 |
| Transport of small molecules | 0.01 | 0.04 |
| HDL clearance | 0.01 | 0.04 |
| Intestinal absorption | 0.01 | 0.04 |
| Scavenging by Class B Receptors | 0.02 | 0.05 |
| GDP-fucose biosynthesis | 0.02 | 0.05 |
| HDL assembly | 0.02 | 0.05 |
| Nuclear Receptor transcription pathway | 0.02 | 0.05 |
| Transport of bile salts and organic acids, metal ions and amine compounds | 0.02 | 0.05 |
| RUNX2 regulates genes involved in cell migration | 0.02 | 0.05 |
| Chylomicron remodeling | 0.03 | 0.05 |
| Chylomicron assembly | 0.03 | 0.05 |
| Zinc influx into cells by the SLC39 gene family | 0.03 | 0.05 |
| HDL remodeling | 0.03 | 0.05 |
| Mucopolysaccharidoses | 0.03 | 0.05 |
| Apoptotic cleavage of cell adhesion proteins | 0.03 | 0.05 |
| SLC-mediated transmembrane transport | 0.03 | 0.05 |
| Hyaluronan uptake and degradation | 0.03 | 0.05 |
| Metabolism of proteins | 0.03 | 0.05 |
| Platelet activation, signaling and aggregation | 0.03 | 0.05 |
| PPARA activates gene expression | 0.04 | 0.05 |
| Signaling by Receptor Tyrosine Kinases | 0.04 | 0.05 |
| Regulation of lipid metabolism by Peroxisome proliferator-activated receptor $\alpha$ | 0.04 | 0.05 |
| Hyaluronan metabolism | 0.04 | 0.05 |
| Vitamin B5 (pantothenate) metabolism | 0.04 | 0.05 |
| Zinc transporters | 0.04 | 0.05 |
| ABC transporters in lipid homeostasis | 0.05 | 0.05 |
| Metabolism of Angiotensinogen to Angiotensins | 0.05 | 0.05 |
| Plasma lipoprotein assembly | 0.05 | 0.05 |
| Scavenging by Class A Receptors | 0.05 | 0.05 |

The pathway analysis report was generated by Reactome (v70) for species "Homo sapiens". These significant data (with a $p$ / FDR <0.05) are based on the overrepresentation analysis (hypergeometric distribution).

**Table 4A.** Down-regulated proteins: Unbiased proteomic screen.

| Proteins | <u>Aquamin</u> |  |  |  |
| --- | --- | --- | --- | --- |
|  | 1.5 mM | 2.1 mM | 3.0mM | 4.5mM |
| Neurogranin | *0.54±0.05 | *0.38±0.16 | *0.30±0.13 | *0.18±0.09 |
| Fibrinogen silencer-binding protein<sup>#</sup> | *0.61±0.09 | *0.44±0.10 | *0.36±0.03 | *0.22±0.13 |
| Stathmin | *0.58±0.07 | *0.47±0.02 | *0.37±0.01 | *0.29±0.12 |
| DNA replication licensing factor MCM3 | *0.70±0.07 | *0.57±0.06 | *0.44±0.07 | *0.34±0.13 |
| Nucleolar GTP-binding protein 2 | *0.59±0.01 | *0.64±0.15 | *0.55±0.04 | *0.30±0.08 |
| Heterogeneous nuclear ribonucleoprotein A0 | *0.71±0.09 | *0.55±0.19 | *0.49±0.20 | *0.39±0.16 |
| Pumilio homolog 3<sup>#</sup> | 0.78±0.25 | *0.70±0.06 | *0.46±0.14 | *0.33±0.19 |
| U3 small nucleolar RNA-interacting protein 2 | *0.62±0.03 | *0.51±0.06 | *0.38±0.01 | *0.32±0.10 |
| Ribonucleoside-diphosphate reductase | *0.68±0.06 | *0.56±0.21 | *0.46±0.14 | *0.35±0.15 |
| Inosine-5'-monophosphate dehydrogenase 2 | *0.63±0.03 | *0.56±0.13 | *0.43±0.11 | *0.35±0.12 |
| DNA replication licensing factor MCM2 | *0.70±0.06 | *0.62±0.16 | *0.47±0.15 | *0.32±0.11 |
| Proliferating cell nuclear antigen | *0.65±0.01 | *0.59±0.08 | *0.45±0.05 | *0.36±0.10 |
| Proliferation marker protein Ki-67 | *0.58±0.08 | *0.48±0.11 | *0.39±0.13 | *0.31±0.08 |
| Nucleolar and coiled-body phosphoprotein 1 | *0.63±0.05 | *0.53±0.06 | *0.44±0.06 | *0.37±0.10 |
| Nucleophosmin | *0.59±0.06 | *0.55±0.06 | *0.43±0.04 | *0.35±0.09 |
| Nuclear autoantigenic sperm protein | *0.66±0.07 | *0.57±0.03 | *0.45±0.03 | *0.37±0.11 |
| Serine/arginine-rich splicing factor 6 | *0.67±0.15 | *0.56±0.11 | *0.47±0.10 | *0.37±0.10 |
| Serine/threonine-protein kinase VRK1 | *0.70±0.07 | *0.61±0.11 | *0.52±0.11 | *0.41±0.12 |
| Protein RRP5 (NF-kappa-B-binding protein) | *0.72±0.03 | *0.54±0.12 | *0.44±0.15 | *0.41±0.06 |
| Nucleolin | *0.70±0.09 | *0.57±0.02 | *0.48±0.04 | *0.41±0.11 |
| PC4 and SFRS1-interacting protein | *0.72±0.10 | *0.56±0.05 | *0.48±0.05 | *0.41±0.12 |
| Non-histone chromosomal protein HMG-17 | *0.73±0.05 | *0.55±0.18 | *0.49±0.16 | *0.43±0.13 |
| DNA replication licensing factor MCM7 | *0.77±0.08 | 0.74±0.23 | *0.56±0.23 | *0.41±0.12 |
| Heterogeneous nuclear ribonucleoprotein D-like | *0.69±0.06 | *0.59±0.02 | *0.50±0.04 | *0.42±0.11 |
| SWI/SNF complex subunit SMARCC1 | *0.69±0.08 | *0.54±0.07 | *0.48±0.04 | *0.41±0.13 |
| NOP2 | *0.74±0.06 | *0.60±0.09 | *0.51±0.06 | *0.43±0.12 |
| Nucleolar RNA helicase 2 | *0.63±0.07 | *0.52±0.12 | *0.43±0.06 | *0.34±0.04 |
| PIN4 | *0.66±0.09 | *0.61±0.07 | *0.51±0.09 | *0.47±0.14 |
| DNA replication licensing factor MCM5 | *0.70±0.10 | *0.57±0.18 | *0.49±0.16 | *0.40±0.13 |
| CD44 antigen | *0.61±0.01 | *0.58±0.12 | *0.48±0.12 | *0.40±0.09 |
| Cytokine-induced protein with coiled-coil domain | *0.69±0.02 | *0.52±0.13 | *0.45±0.10 | *0.42±0.11 |
| Gem-associated protein 5 | *0.82±0.11 | *0.65±0.13 | *0.53±0.16 | *0.47±0.11 |
| Zinc finger C2HC domain-containing protein 1A | *0.54±0.17 | *0.50±0.09 | *0.65±0.18 | *0.54±0.11 |
| Translation machinery-associated protein 7 | *0.63±0.07 | *0.57±0.10 | *0.49±0.06 | *0.44±0.09 |
| DNA replication licensing factor MCM4 | *0.70±0.02 | *0.67±0.09 | *0.52±0.07 | *0.43±0.10 |
| Ras GTPase-activating protein-binding protein 1 | *0.75±0.07 | *0.61±0.10 | *0.55±0.09 | *0.47±0.10 |
| CD99 antigen | *0.78±0.03 | *0.64±0.10 | *0.59±0.10 | *0.39±0.04 |
| NHP2-like protein 1 | *0.69±0.04 | *0.57±0.08 | *0.53±0.08 | *0.47±0.11 |
| Receptor-type tyrosine-protein phosphatase F | *0.56±0.09 | *0.55±0.11 | *0.50±0.08 | *0.45±0.07 |
| Putative RNA-binding protein Luc7-like 1 | *0.69±0.07 | *0.72±0.11 | *0.58±0.15 | *0.47±0.07 |
| HEAT repeat-containing protein 1 | *0.69±0.17 | *0.64±0.08 | *0.55±0.07 | *0.43±0.03 |
| U6 snRNA-associated Sm-like protein LSm5 | *0.77±0.09 | *0.61±0.12 | *0.56±0.07 | *0.48±0.10 |
| Importin subunit alpha-1 | *0.70±0.08 | *0.66±0.11 | *0.57±0.05 | *0.48±0.04 |
| Palmitoyl-protein thioesterase 1 | *0.66±0.03 | *0.63±0.05 | *0.60±0.03 | *0.49±0.05 |
| Leydig cell tumor 10 kDa protein homolog | *0.67±0.01 | *0.63±0.02 | *0.56±0.02 | *0.49±0.07 |
| Ribosome biogenesis protein WDR12 | *0.77±0.11 | *0.67±0.10 | *0.58±0.04 | *0.47±0.05 |
| Transformer-2 protein homolog beta | 0.85±0.24 | *0.56±0.07 | *0.53±0.04 | *0.48±0.06 |
| Ribosome biogenesis protein BMS1 homolog | 0.92±0.32 | 0.79±0.34 | *0.61±0.15 | *0.49±0.04 |
| Drebrin | *0.70±0.08 | *0.60±0.05 | *0.59±0.02 | *0.53±0.09 |
| BRCA2 and CDKN1A-interacting protein | *0.70±0.06 | *0.68±0.07 | *0.57±0.03 | *0.52±0.06 |
| Ubiquitin-associated protein 2-like | *0.72±0.05 | *0.62±0.07 | *0.57±0.04 | *0.55±0.07 |

The numerical values represent mean ±SD (fold-change of abundance ratio) as compared to the control, based on UC colonic tissue (n=3 subjects). Each subject was assessed separately using TMT based differential proteomic expression and the data from the individual sets averaged together. False Discovery rate (FDR) was <1% except two proteins<sup>#</sup> with FDR<2%. Values were ordered from minimum to maximum fold-change based on the minimum response with any of the Aquamin® concentrations. To be included, values were down-regulated by at least 1.8-fold with at least one concentration of Aquamin® as compared to control and common in all three subjects. Asterisks indicate statistical significance as compared to control at $p < 0.05$ . NOP2: Probable 28S rRNA (cytosine(4447)-C(5))-methyltransferase; PIN4: Peptidyl-prolyl cis-trans isomerase NIMA-interacting 4.

**Table 4B.**
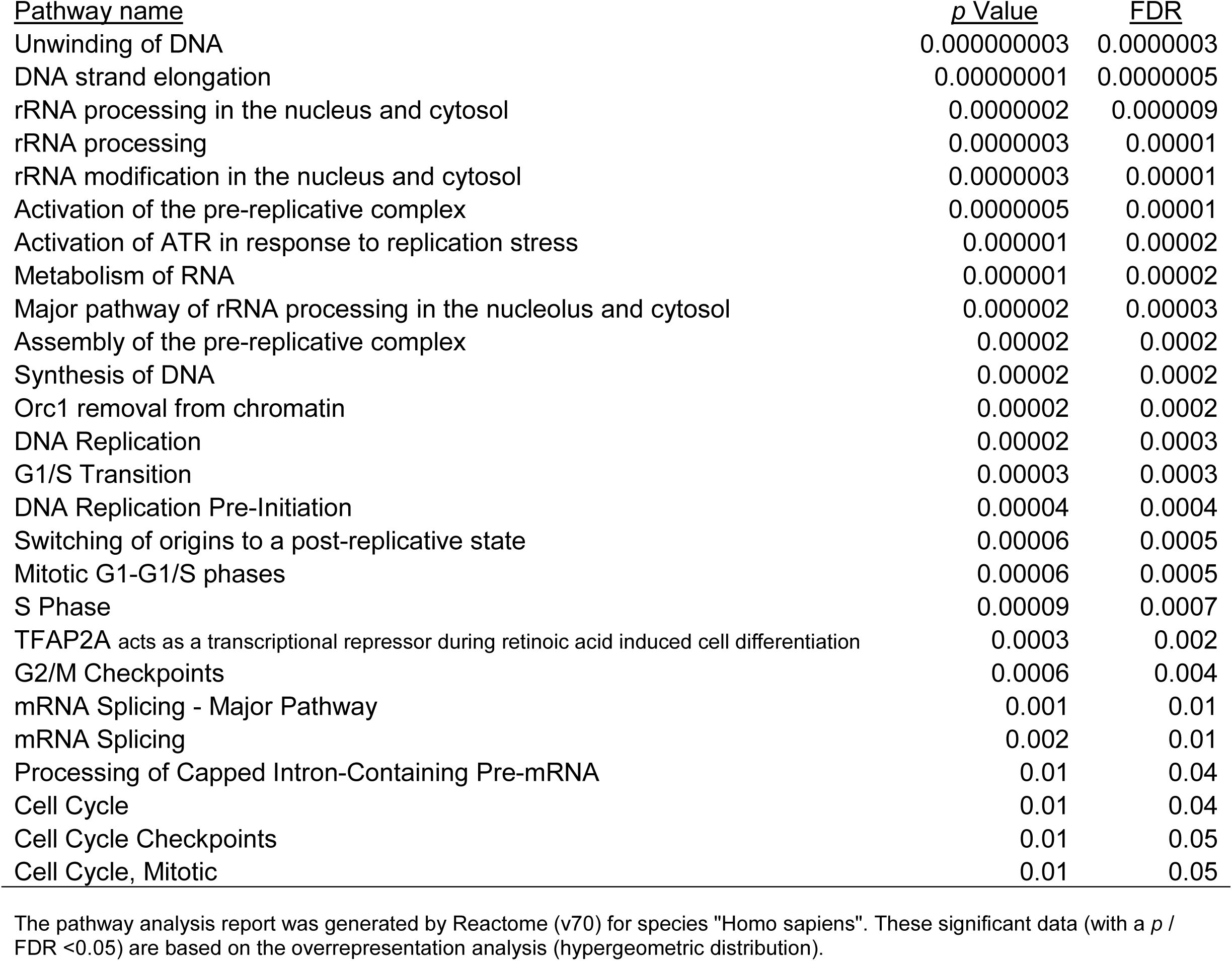
Pathways significantly activated based on down-regulated Proteins.

### Cytokine profile in control and mineral-treated colonoids

In a final set of experiments, we assessed 2-day culture fluids obtained from control and Aquamin^®^-treated colonoid cultures at day-14 (immediately prior to harvest) for levels of pro-inflammatory cytokines (TNF-α, IL-1β, IL6, IL-8 and MCP-1). While levels of TNF-α, IL-1β, IL6 and MCP-1 were near or below quantifiable limits, substantial amounts of IL-8 were detected in all culture fluids. The average level of IL-8 in control specimens was 410 ± 193 pg/mL (n=7) and this was reduced to 362 ± 89 pg/mL and 344 ± 213 pg/mL in specimens treated with Aquamin^®^ (providing 1.5 and 2.1 mM calcium, respectively). For comparison, the level of IL-8 seen in colonoids initiated with tissue from healthy control subjects (available from our previous study [10]) was 268 ± 103 pg/mL (n=3).

## DISCUSSION

The colonic barrier is compromised in UC patients (2-5), but whether this is simply the result of inflammatory injury to the epithelial lining, or whether pre-existing barrier weakness is a contributor to disease susceptibility has not been fully resolved. Regardless, it seems reasonable to suggest that strengthening the mucosal barrier could be of value. At the very least, an intact barrier would limit the permeation of both soluble and particulate pro-inflammatory moieties from the gut into the interstitium. Improvement in the barrier, especially if it involved the basement membrane, might also retard passage of inflammatory cells into the epithelium to, presumably, reduce epithelial cell injury. The potential to improve the colonic barrier, as much as anything, drives the search for natural products and other adjuvant therapies for use in individuals with UC in remission. Such therapies are unlikely to replace the potent anti-inflammatory drugs used in the treatment of acute disease flare-ups. None-the-less, improvement in barrier function in UC patients who are in remission may be of value to many individuals in so far as this could help maintain the state of remission and reduce symptomatology during this period. The findings presented in this manuscript suggest that an appropriate level of mineral intake may be of value in this regard.

Here we demonstrate that treatment of UC tissue in colonoid culture with Aquamin^®^, a natural product consisting of calcium and magnesium along with 72 additional trace elements, up-regulates multiple proteins that contribute to the formation of a healthy barrier in the colon. Among these are proteins that comprise the primary cell-cell adhesion complexes – tight junctions, adherens junctions and, especially, desmosomes. Also up-regulated are major non-collagenous components of the basement membrane along with hemidesmosomal components. Factors present on the apical surface (mucins, trefoils and CEACAMS) are also up-regulated. Which of the multiple changes observed here are most important to improved barrier formation remains to be determined. While most studies have focused on tight junctions and on desmosomes (21-24), other cellular and extracellular structures contribute. Of these, the basement membrane may be the most important (25-28). The basement membrane supports barrier function in several important ways. The basement membrane inhibits bacterial crossing from the colonic stream into the interstitium and limits leukocyte trafficking from the circulation into the colonic epithelium. Additionally, the basement membrane is rich in anionic substances (heparin sulfate proteoglycans) – providing capacity to trap cationic moieties and regulate their permeation. Perhaps most importantly, the colonic wall consists of a single layer of epithelial cells and each cell is attached to the basement membrane. When cells detach from this substratum, they rapidly undergo apoptosis. With that, all barrier function is compromised. Of interest in this regard, a loss of laminin in the colonic wall of individuals with UC has been noted (29,30). Also of interest, past studies have shown that disruption of the basement membrane or interference with epithelial cell binding to laminin (the major non-collagenous glycoprotein component of the basement membrane) results in widespread colitis in mice (31).

Proteins expressed at the apical surface (mucins and trefoils) also have a role to play. Mucins secreted by goblet cells are carbohydrate-rich molecules (32). Trefoils are a family of proteins that interact with mucins and help organize the mucins into a viscous mucous network to trap bacteria and other particulates and to prevent their reaching the cell surface. Disruption of this network has been linked to inflammatory bowel disease (20,21).

Along with structural components of the barrier, a number of peptides with known anti-microbial activity were induced in response to the multi-mineral intervention (Table 2). These too could be considered part of the barrier, as could the bacterial film present on the colonocyte surface and bacteria present in the fecal stream (33). When a healthy microbial community is present, it limits the overgrowth of potentially harmful microbes. In part, this occurs via the healthy bacteria simply occupying a niche that could otherwise be colonized by pathogens. At the same time, the healthy microbial population can contribute to the barrier through elaboration of beneficial metabolites (e.g., short chain fatty acids [SCFA]) rather than toxic bile acids. While colonoid culture does not provide a means to assess changes in the gut microbial community, we are (in parallel with the current study) conducting a 90-day interventional trial with Aquamin^®^ in healthy human subjects Clinicaltrials.gov; NCT02647671). In findings to date, Aquamin^®^ ingestion altered the colonic microbial profile and, concomitantly, decreased bile acids levels and increased the level of the major SCFA, acetate, as compared to placebo (or calcium alone) (34). While the ongoing interventional trial is being conducted with healthy volunteers, a similar study in patients with UC in remission has just begun (Clinicaltrials.gov; NCT03869905).

Finally, our proteomic screen identified Aquamin^®^-induced changes in a number of proteins linked to gut inflammation. Pro-inflammatory elements were uniformly down-regulated while moieties with anti-inflammatory, anti-oxidant and anti-apoptotic activity were increased. It should be noted in regard to the inflammation-modulating moieties that these were detected in the colonic tissue, itself, where cellular injury occurs. Of the secreted moieties related to inflammation, only IL-8 was detected in large amounts; it was decreased with Aquamin^®^.

Growth-reduction accompanies differentiation in epithelial cells (18). While our unbiased proteomic analysis showed that most of the up-regulated proteins were part of the differentiation process, many of the down-regulated proteins were related to proliferation. Consistent with this, a reduction in Ki67 expression with intervention was seen by immunostaining. It could be argued that suppression of epithelial cell proliferation in the context of a colonic ulcer might be an unwanted consequence of a differentiation-promoting intervention. While this possibility needs to be acknowledged, it is unlikely that effects on proliferation would counteract the beneficial activities related to improved barrier formation and reduced inflammation.

Hyperplastic epithelial growth is part of the wound-healing process, including in the colon (35), but its continued presence is a reflection of chronic wound formation. Thus, differentiation-mediated suppression of hyperplastic growth in the epithelium (occurring as a normal component of the wound-healing process) is, ultimately, necessary. Furthermore, colonic hyperplasia / dysplasia is prognostically-related to cancer development in UC patients (31,36). A recent study (36) has demonstrated that the accumulation of a protein referred to as Stathmin (STMN1) in UC lesions is associated with neoplasia. STMN1 was significantly down-regulated by Aquamin^®^ (up to 3.5 fold-reduction; Table 4A). Thus, proliferation-regulating effects of Aquamin^®^ might contribute to reduced cancer development in UC patients. Ultimately, these complex questions will need to be addressed experimentally, and this is beyond the scope of the current work.

## CONCLUSIONS (and implications)

A schematic representation of the colonic mucosa in UC-derived colonoid culture is presented in Figure 3. It highlights structural changes in the barrier occurring in response to the multi-mineral intervention. Based on the results presented here, we hypothesize greater tissue strength, improved cell-basement membrane interaction and a thicker mucous layer above the apical surface. These changes along with the increased elaboration of anti-microbial and anti-inflammatory proteins should, ultimately, be beneficial to individuals with UC in remission.

**Figure 3.**
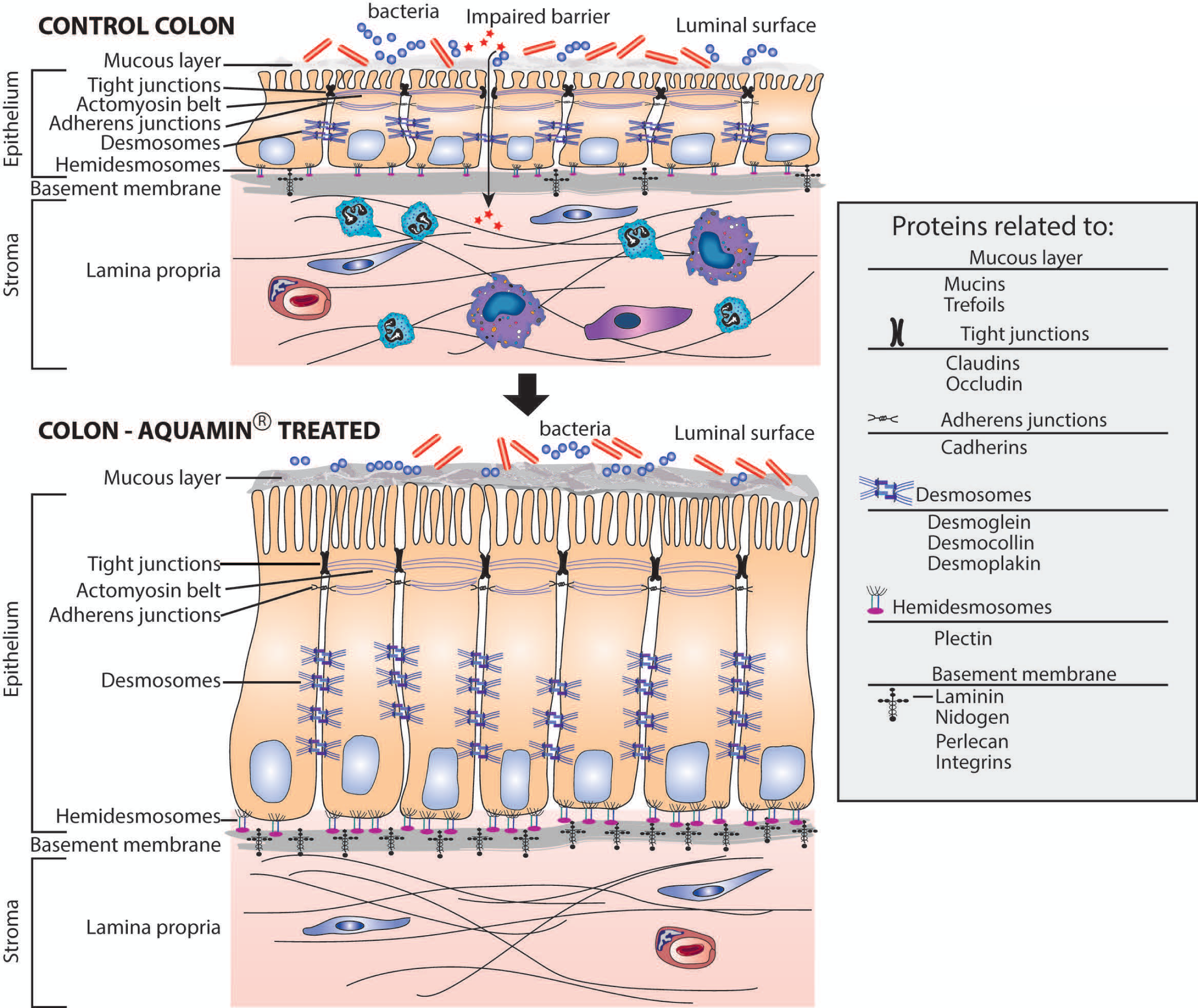
**Schematic representation of the colonic mucosa in UC-derived colonoid culture and structural changes due to intervention with Aquamin^®^.** Tight junctions are observed at the apical surface between adjacent cells in both control and treated colonoids; there is little observable difference between the two. Desmosomes (shown along the lateral surface between cells) are increased in response to treatment. This should support increased tissue strength. Additional changes resulting from Aquamin^®^ intervention include an increase in the non-collagenous components of the basement membrane and an increase in hemidesmosomal proteins. These changes should promote improved cell-matrix adhesion. Increased mucin and trefoil levels, leading to a thicker mucous layer at the luminal surface should contribute to more efficient trapping of bacteria. In aggregate, these changes should provide for improved barrier function and (hopefully) help mitigate colonic inflammation.

A majority of individuals in Western society do not reach the recommended daily intake for calcium (according to U.S. Department of Agriculture. 2015 – 2020 Dietary Guidelines for Americans; available at https://health.gov/dietaryguidelines/2015/guidelines/), and this deficiency contributes to multiple chronic diseases (37,38). Magnesium-deficiency is also common (39). While there are no intake recommendations for many of the other potentially important trace elements in Aquamin^®^, deficiencies likely exist for those elements that are nutritionally associated with calcium or magnesium. While roles for calcium and magnesium in prevention of chronic disease are, at least partially, understood, what role micronutrient deficiencies – including those involving minerals – play in chronic or long-latency diseases is only beginning to get attention (39,40). To the extent that multi-mineral deficiencies are common, the findings presented here allow us to hypothesize that such deficiencies might contribute to weakening of the colonic barrier in many individuals, including those with UC. Obtaining the proper intake of minerals, either through a healthful eating pattern or through the use of an appropriate multi-mineral supplement, should be beneficial.

Whether the multi-mineral product investigated here (Aquamin^®^) or some other mineral formulation will, ultimately, provide benefit to individuals with UC and whether the intervention will prove safe and tolerable needs to be established through precisely controlled clinical studies. While the necessary studies have not yet been conducted, we have just begun a pilot-phase trial (Clinicaltrials.gov; NCT03869905) with UC patients in remission to address safety and tolerability concerns, and to begin generating efficacy data with Aquamin^®^.

## Data Availability

All the data are fully available within the supplementary files.

## ACKNOWLEDGEMENTS

We thank Marigot LTD (Cork, Ireland) for providing Aquamin^®^ as a gift. We thank the Proteomic Resource Facility (Pathology Department) for help with proteomic data acquisition; the Histology and Immunohistology Laboratory (University of Michigan Comprehensive Cancer Center) for immunohistochemical staining; the Microscopy and Imaging Laboratory (MIL) for help with electron microscopy and the Slide-Scanning Services (Pathology Department) for assistance with slide scanning. We thank the Translational Tissue Modeling Laboratory (TTML) at University of Michigan for help with ulcerative colitis-derived colonoid initiation and propagation. Finally, our thanks go to Ron Allen for help with cytokine assessment.

## SUPPLEMENT FIGURE LEGENDS

**Supplement Figure 1. Ki67 and cadherin-17 expression by immunohistology**. At the end of the incubation period, colonoids were examined after immunostaining of histological sections. Ki67 (**a-e**); Bar=200µm and Cadherin-17 **(f-j);** Bar=100µm. Quantitative assessment of Ki67 staining (**k**) is based on nuclear algorithm (v9) and data represent means and standard deviations from n=3 subjects and 36-78 individual crypts per condition. Asterisks (*) indicate statistical significance from control at p<0.05. Cadherin-17 (**l**) values represent positivity (measured using Positive Pixel Value v9). Means and standard deviations are based on n=3 subjects and 68-124 individual crypts per condition. Asterisks (*) indicate statistical significance from control at p<0.05. CDH17: Cadherin-17

**Supplement Figure 2. Protein distribution with differing Aquamin**^**®**^ **levels across three subjects**. At the end of the incubation period, lysates were prepared for proteomic analysis. **Upper panel**: Venn plots (generated by BioVenn, a visualization tool) showing the number of proteins altered (increased or decreased) by an average of 1.8-fold or greater in each of the three data sets and the overlap across the three specimens at each concentration of Aquamin^®^. **Lower panel**: Venn plot showing distribution of the common proteins (from the top panels) altered (increased or decreased) by each concentration of Aquamin^®^ with an average of 1.8-fold or greater and each individual specimen. These data provide an indication of variability among individual subjects in their response to each concentration of Aquamin^®^. DSG2: Desmoglein-2; CDH17: Cadherin-17; LYPD8: Ly6/PLAUR domain-containing protein 8.

## FIGURES AND TABLES (in body of text)

Table 1. Demographic information

Figure 1. Gross and histological appearance plus immunohistology for CK20

Table 2. Proteomic Table – Differentiation and barrier molecules (plus anti-microbial and inflammation-related)

Figure 2. Immunohistology and TEM – desmoglein-2 and desmosomes

Table 3. Up-regulated proteins and pathways – unbiased search

Table 4. Down-regulated proteins and pathways – unbiased search

Figure 3. Barrier cartoon

## SUPPLEMENTAL MATERIAL

Supplement Table 1. Mineral composition of Aquamin^®^ Supplement Table 2. Antibody related information

Supplement Table 3. A list of altered (and common) proteins in response to different concentrations of Aquamin^®^ at 1.8 fold-change.

Supplement Table 4. Comprehensive list of altered proteins with Aquamin^®^ (*p* value <0.05 across all conditions) Supplement Figure 1. IHC for Ki67 and cadherin-17

Supplement Figure 2. Relationship of protein changes comparing proteomic signature of three UC patients.

Supplemental Text File 1. Relevant citations to differentiation / barrier related proteins presented in the Table 2.

### ABBREVIATIONS

UC: Ulcerative colitis
SEM: Scanning electron microscopy
TEM: Transmission electron microscopy
TEER: Trans-epithelial electrical resistance
TGF: Transforming Growth Factor
ROCK: Rho-associated, coiled-coil containing protein kinase
EDTA: Ethylenediaminetetraacetic acid
TRS: Target Retrieval Solution
HRP: Horseradish peroxidase
FDR: False discovery rate
ANOVA: Analysis of variance
SD: Standard deviation
CK20: Cytokeratin 20
CDH17: Cadherin-17
DSG2: Desmoglein-2
CEACAM: Carcinoembryonic antigen-related cell adhesion molecule
SCFA: Short-chain fatty acids
TNF-α: Tumor necrosis factor-alpha
IL: Interleukin
MCP-1: Macrophage chemotactic peptide-1

